# Willingness to Enroll in Social Health Insurance and associated factors among Household Heads in Obio/Akpor Local Government Area of Rivers State

**DOI:** 10.1101/2022.12.19.22283656

**Authors:** Tabansi Chinelo Kenechukwu, Tamunotonye Harry, Uchechukwu Apugo

## Abstract

**Background:** Social health insurance (SHI) schemes cover the healthcare needs of the informal sector workers, provide them with financial protection, minimize the equity gap, reduce Out-Of-Pocket (OOP) spending, and enhance utilization of the healthcare system.

**Objective:** To determine the willingness to enrol in SHI and its associated factors among household heads in Obio-Akpor Local Government Area (LGA) of Rivers state.

**Method:** This community-based cross-sectional study was carried out using the clusters and the EPI random walk sampling technique to obtain data from 205 selected household heads who were administered with an interviewer-administered questionnaire. The collected data was analysed using SPSS version 21.

**Result:** According to the findings, majority of the respondents were males (58.5%), aged between 31 – 45 years (52.7%), married (57.1%), completed the tertiary level of education (65.4%), employed or self-employed (95.1%), from a family size of 3 – 4 (56.1%) and earned 50,000 to 100,000 Naira monthly (27.8%). Also, 63.0% are not enrolled in any health insurance scheme out of which 60.5% are not willing to enrol, with lack of trust in the management of the scheme, no believe in paying for sickness or have other means of meeting their healthcare needs identified as the major reasons. However, younger age, higher educational level, employment status, and earning >50,000 – 100,000 monthly had statistically (p<0.05) significant effect on the willingness of the respondents to enrol on SHI

**Conclusion:** This result showed a low level of enrolment in health insurance as well as willingness to enrol in Shi, hence, both insurance companies, government and even Non-Governmental Organisation should carry out strategic campaigns to dispel false rumours about SHI while outlining the merits.

## INTRODUCTION

Providing healthcare, till date, still remains one of the most essential basic social services that any government or organization can provide to its citizens (Aderibigbe et al., 2017; Azuogu & Eze, 2018). However, due to inadequate public investment in healthcare delivery, residents in developing nations, particularly in Sub-Saharan Africa and Asia are confronted with a variety of problems in their quest for quality healthcare. This encourages the majority of people in these countries to go from using public healthcare to seeking out alternative medicine, which is regarded to be less expensive. According to the statistics found in the Nigerian National Health Financing Policy, roughly 4% of households spend more than half of their whole household expenditures on healthcare, and 12% spend more than a quarter of their total household expenditures on healthcare (Ezenekwe et al., 2020). Also, in contrast to adopting the use of health insurance programs, which would have made the service cheap, affordable, and reliable, the healthcare system in these nations mostly employ out-of-pocket payment (OOP) as a method of payment for healthcare services. According to Alo, Okedo-Alex, and Akamike (2020), roughly 100 million individuals become impoverished each year as a result of OOP health expenditures, with another 1.2 billion becoming poorer as a result of the same OOP health spending. Inevitably, this type of healthcare finance can lead to unfairness and poor health-seeking behaviour (Goudge et al., 2009) and greatly hinders the achievement of Universal health coverage (UHC) which is aimed at safeguarding people from financial hardship owing to catastrophic health spending (WHO, 2012).

As stated by Anderson and Adeniji (2019), the finance pattern of a country’s healthcare system is a determining factor in achieving UHC, while the most viable strategy to achieving UHC is through the provision of subsidized and cheap health insurance coverage, which generally includes protection against the risk of incurring personal medical costs (Alesane & Anang, 2018). Hence, governments in many developing countries have resorted to constructing health insurance models to address these difficulties in order to avoid catastrophic health costs and the necessity to strive for UHC (Fox & Reich, 2015; Kusi et al., 2015; Badu et al., 2018). In Nigeria, the Federal Government officially launched the National Health Insurance Scheme (NHIS) on the 6th of June 2005, with its participation across the country voluntary but mandatory for public civil servants in Federal Establishments who have been the major beneficiaries of this scheme since its inception (Akande, Salaudeen & Babatunde, 2011). Hence, reports show that it only has about 5% enrolment because it is largely limited to government employers and formal sector workers with regular jobs, leaving the unemployed without coverage (Odeyemi & Nixon, 2013; Shafie & Hassali, 2013). This buttresses the need for the adoption of a social health insurance (SHI) scheme to cover the healthcare needs of the informal sector workers, provide them with financial protection, minimize the equity gap and reduce OOP spending, and enhance utilization of the health care system (Anderson & Adeniji, 2019). However, though the utilisation of SH has been associated with increased access and utilisation of healthcare services in most developed nations (Kusi et al., 2015), such schemes have one or more issues in the eyes of the users, as most of these plans, according to Jain et al. (2014), do not cover entire healthcare services but only secondary and tertiary care hospitalization charges. This, among other reasons, has influenced people’s desire to enrol in any insurance system, even at the community level, because they still have to pay to use the hospital’s services because what their insurance covers may not be adequate. Hence, this study was geared towards determining the willingness to enrol in social health insurance and its associated factors among household heads in Obio-Akpor Local Government Area (LGA) of Rivers state.

## METHODS

### Study setting and Design

This study was designed as a community-based cross-sectional study which was carried out in Obio-Akpor Local Government Area (LGA). The LGA is one of the 23 LGAs in Rivers state and one of the two urban LGAs in the state, as well as one of the major centres of economic activities in Nigeria. According to the All Answers Limited (2018), the LGA has 20 Primary Health Care centres, 12 are comprehensive health care centres, 3 are primary health care clinics and 5 are basic health clinics of which 3 are health posts. Also, there are a few secondary health care facilities and 1 tertiary hospital which is the University of Port Harcourt Teaching Hospital (UPTH).

### Study Population and Sample size

The study population was derived from residents of the communities in Obio-Akpor LGA (OBALGA) who head of their families or any responsible member of the house. The sample size of 204 household heads was derived by applying the Cochrane single population proportion formula (N = pqz^2^/d^2^), and considering; 95% confidence interval, 5% margin of error, 86% proportion of individuals willing to participate in health insurance scheme (Anderson & Adeniji, 2019), and a 10% non-response rate. The sampling method involved a two-stage sampling, using firstly wards (clusters) and the EPI random walk method (WHO, 2008; Russo et al., 2015) for the selection of 12 households from 17 selected communities in the LGA, due to the unavailability of a comprehensive list of households.

### Study Instrument

Data for the study was collected with the aid of a pre-tested, semi-structured, interviewer-administered questionnaire which was adapted from the WHO World Health Survey Tool as used in the study of Yusuf et al. (2019). It is made up of two sections which collected information on socio-demographic characteristics of the respondents and their willingness to enrol in a social health insurance scheme.

### Data Analysis

The data from the study were entered into Excel Spreadsheet, cleaned and statistical analysis done using IBM SPSS version 21. Frequencies and proportions were calculated and tabulated, while Chi-square statistics was used to determine the relationship between willingness to enrol for SHI and selected socio-demographic characteristics of the respondents, with the level of significance set as P < 0.05.

### Ethical Consideration

The study was approved by the Research and Ethics Committee of the University of Port Harcourt while official permission was obtained from the community heads and written consent from the household heads.

## RESULT

The socio-demographic and economic characteristics of the respondents recruited for this study from the communities in OBALGA is presented in table 1 above. According to the result, a total of 205 household heads were recruited in the study, out of which majority were males (58.5%), aged between 31 – 45 years (52.7%), married (57.1%), completed the tertiary level of education (65.4%), employed or self-employed (95.1%), from a family size of 3 – 4 (56.1%) and earned 50,000 to 100,000 Naira monthly (27.8%).

**Table 1:**
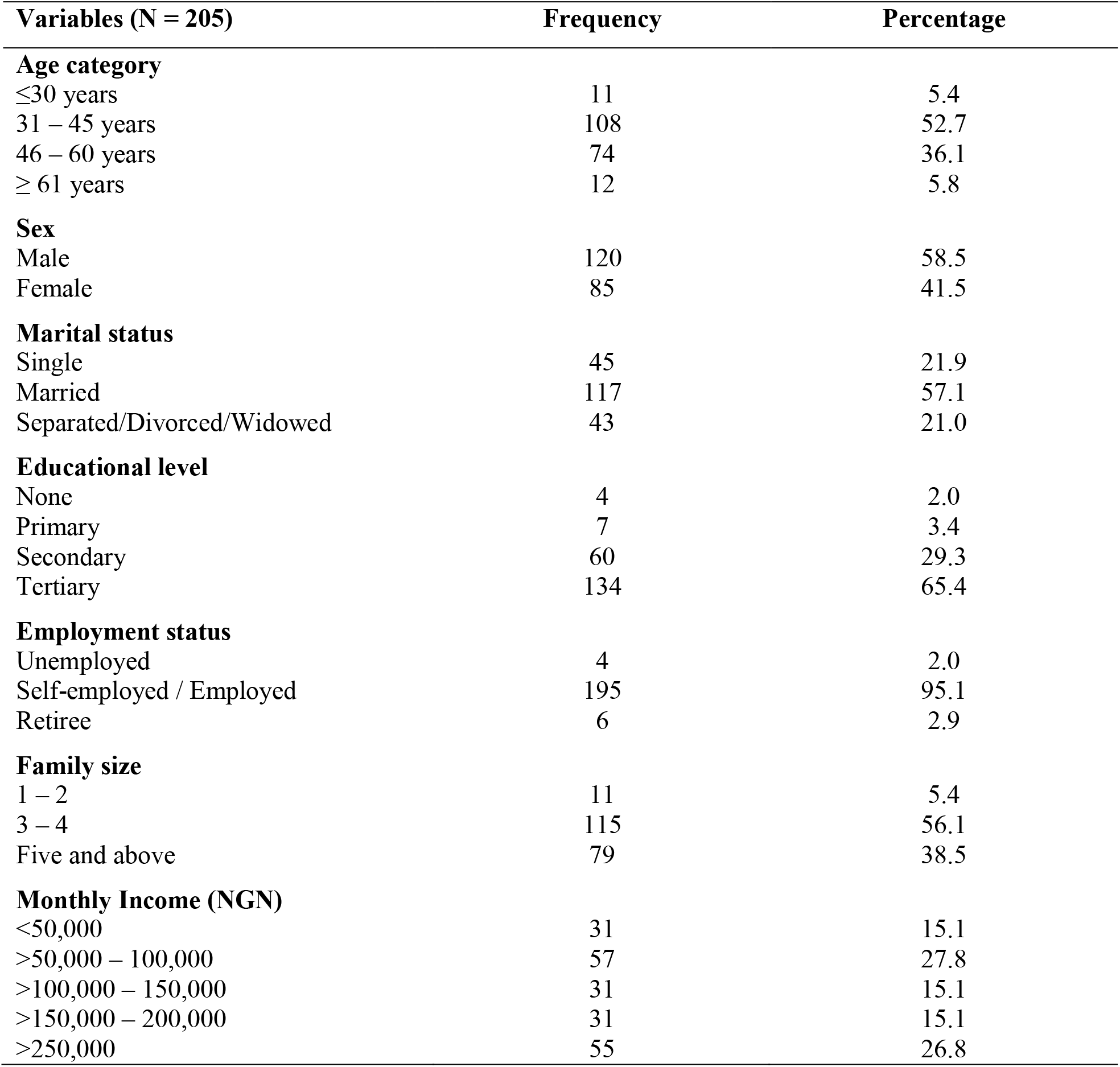
Socio-demographic characteristics of household heads in Obio-Akpor LGA

| Variables (N = 205) | Frequency | Percentage |
| --- | --- | --- |
| **Age category** |  |  |
| ≤30 years | 11 | 5.4 |
| 31 – 45 years | 108 | 52.7 |
| 46 – 60 years | 74 | 36.1 |
| ≥ 61 years | 12 | 5.8 |
| **Sex** |  |  |
| Male | 120 | 58.5 |
| Female | 85 | 41.5 |
| **Marital status** |  |  |
| Single | 45 | 21.9 |
| Married | 117 | 57.1 |
| Separated/Divorced/Widowed | 43 | 21.0 |
| **Educational level** |  |  |
| None | 4 | 2.0 |
| Primary | 7 | 3.4 |
| Secondary | 60 | 29.3 |
| Tertiary | 134 | 65.4 |
| **Employment status** |  |  |
| Unemployed | 4 | 2.0 |
| Self-employed / Employed | 195 | 95.1 |
| Retiree | 6 | 2.9 |
| **Family size** |  |  |
| 1 – 2 | 11 | 5.4 |
| 3 – 4 | 115 | 56.1 |
| Five and above | 79 | 38.5 |
| **Monthly Income (NGN)** |  |  |
| <50,000 | 31 | 15.1 |
| >50,000 – 100,000 | 57 | 27.8 |
| >100,000 – 150,000 | 31 | 15.1 |
| >150,000 – 200,000 | 31 | 15.1 |
| >250,000 | 55 | 26.8 |

### Willingness to enrol in social health insurance

The result as tabulated in table 2 above revealed that over half of the respondents (63.0%) are not enrolled in any health insurance scheme. Among the 129 respondents that are currently not enrolled in any health insurance scheme, 78 (60.5%) of them responded that they are not willing to enrol in a SHI while 51 (39.5%) expressed their willingness. Among the 51 household heads that declared interest to enrol in a SHI scheme, majority (78.4%) stated that they would like to enrol in a privately managed scheme while those who wanted the hospital and government-based scheme formed the remaining 13.7% and 7.9% respectively. When asked how much they are willing to pay annually for a SHI scheme, majority (70.6%) reported that they can contribute 10,000 – 50,000 Naira towards the scheme. Among those who declared their unwillingness to enrol in SHI, a greater majority (43.6%) stated that they are not just interested while 20.5% stated that they do not just believe in paying for sickness. 12.8% are not interested because they have other means of healthcare while 10.3% are afraid of fund mismanagement. Other issues represented were lack of regular income (6.4%) and lack of trust in the system (3.8%) while only 2(2.6%) of the household heads reported that they do not need social health insurance.

**Table 2:** Willingness to enrol in health insurance scheme among respondents in the study

| Variables | Frequency | Percentage |
| --- | --- | --- |
| **Are you already enrolled or contributing financially to a health insurance scheme** |  |  |
| Not enrolled | 129 | 63.0 |
| Enrolled and renewing | 66 | 32.2 |
| Enrolled but yet to renew | 10 | 4.8 |
| **Willing to enrol in social health insurance scheme (n = 129)** |  |  |
| Yes | 51 | 39.5 |
| No | 78 | 60.5 |
| **Types of health insurance cover like to enroll in (n = 51)** |  |  |
| Private based | 40 | 78.4 |
| Hospital based | 7 | 13.7 |
| Government based | 4 | 7.9 |
| **How much willing to pay annually for health insurance (n = 51)** |  |  |
| < ₦10,000 | 5 | 9.8 |
| ₦ 10,000 – ₦ 50,000 | 36 | 70.6 |
| ₦ 60,000 – ₦ 100,000 | 7 | 13.7 |
| > ₦ 100,000 | 3 | 5.9 |
| **Reasons for unwillingness to enrol in SHI (n=78)** |  |  |
| Not interested | 34 | 43.6 |
| Don't believe in paying for sickness | 16 | 20.5 |
| Have other means of healthcare | 10 | 12.8 |
| Afraid of fund mismanagement | 8 | 10.3 |
| Lack of regular income | 5 | 6.4 |
| Don't trust the system will work | 3 | 3.8 |
| Don't need health insurance | 2 | 2.6 |

### Factors Associated with Willingness to Enrol in Social Health Insurance

The factors associated with the willingness of the respondents towards enrolling for SHII was also analysed and presented in table 3. According to the findings, younger age (≤ 30 years), higher educational level (Tertiary), employment status (Employed), and earning >50,000 – 100,000 monthly had statistically (p<0.05) significant effect on the willingness of the respondents to enrol on SHI.

**Table 3:** Factors associated with willingness to enrol for SHI among the household heads

| Variables (N = 129) | Willing to enrol in SHI schemes |  | Total n(%) | p-value |
| --- | --- | --- | --- | --- |
|  | Yes n(%) | No n(%) |  |  |
| **Age category** |  |  |  |  |
| ≤30 years | 8 (72.7) | 3 (27.3) | 11 (100.0) | **0.002\*** |
| 31 – 40 years | 20 (38.5) | 32 (61.5) | 52 (100.0) |  |
| 41 – 50 years | 10 (23.3) | 33 (76.7) | 43 (100.0) |  |
| 51 – 60 years | 13 (65.0) | 7 (35.0) | 20 (100.0) |  |
| 61 – 70 years | 0 (0.0) | 2 (100.0) | 2 (100.0) |  |
| ≥70 years | 0 (0.0) | 1 (100.0) | 1 (100.0) |  |
| **Sex** |  |  |  |  |
| Male | 31 (40.3) | 46 (59.7) | 77 (100.0) | 0.838 |
| Female | 20 (38.5) | 32 (61.5) | 52 (100.0) |  |
| **Marital status** |  |  |  |  |
| Single | 14 (42.4) | 19 (57.6) | 33 (100.0) | 0.671 |
| Married | 28 (40.0) | 42 (60.0) | 70 (100.0) |  |
| Co-habiting | 0 (0.0) | 2 (100.0) | 2 (100.0) |  |
| Separated/Divorced | 3 (60.0) | 2 (40.0) | 5 (100.0) |  |
| Widowed | 6 (31.6) | 13 (68.4) | 19 (100.0) |  |
| **Educational level** |  |  |  |  |
| None | 1 (33.3) | 2 (66.7) | 3 (100.0) | **0.0001\*** |
| Primary | 2 (28.6) | 5 (71.4) | 7 (100.0) |  |
| Secondary | 11 (20.4) | 43 (79.6) | 54 (100.0) |  |
| Tertiary | 37 (56.9) | 28 (43.1) | 65 (100.0) |  |
| **Employment status** |  |  |  |  |
| Unemployed | 1 (50.0) | 1 (50.0) | 2 (100.0) | **0.0001\*** |
| Employed | 29 (61.7) | 18 (38.3) | 47 (100.0) |  |
| Self-employed | 20 (25.6) | 58 (74.4) | 78 (100.0) |  |
| Retiree | 1 (50.0) | 1 (50.0) | 2 (100.0) |  |
| **Family size** |  |  |  |  |
| One | 1 (33.3) | 2 (66.7) | 3 (100.0) | 0.786 |
| Two | 3 (60.0) | 2 (40.0) | 5 (100.0) |  |
| Three | 12 (40.0) | 18 (60.0) | 30 (100.0) |  |
| Four | 17 (45.9) | 20 (54.1) | 37 (100.0) |  |
| Five or more | 18 (33.3) | 36 (66.7) | 54 (100.0) |  |
| **Monthly Income (₦)** |  |  |  |  |
| <50,000 | 8 (29.6) | 19 (70.4) | 27 (100.0) | **0.026\*** |
| >50,000 – 100,000 | 12 (26.7) | 33 (73.3) | 45 (100.0) |  |
| >100,000 – 150,000 | 10 (52.6) | 9 (47.4) | 19 (100.0) |  |
| >150,000 – 200,000 | 7 (43.8) | 9 (56.2) | 16 (100.0) |  |
| >250,000 | 14 (63.6) | 8 (36.4) | 22 (100.0) |  |

\*Statistically significant ( $p < 0.05$ )

## DISCUSSION

The implementation of health insurance as part of health reform programmes and strategies have been channelled towards providing effective and efficient health care for citizens, most especially for the poor and vulnerable (Azuogu & Eze, 2018). Hence, this community-based study was geared towards ascertaining the level of enrolment into SHI as well as willingness to enrol and its associated factors among household heads in OBALGA.

### Willingness to enrol in social health insurance

Findings from this study show that over half (60.5%) of the respondents are not willing to enrol in a SHI with the major reasons identified as lack of interest and do not believe in paying for sickness while the rest stated that they have other means of meeting their healthcare needs and are afraid of fund mismanagement. This stands as a major setback towards an effective implementation of the scheme as the required resource mobilization and increased size of the risk pool may not be actualized in the community. This low level of the willingness of the household heads in the community in enrolling for SHI agrees with the findings of Tadele et al. (2017) where only 24.1 % of the participants were willing to enrol for SHI. Also, Onwujekwe et al. (2010) in South East Nigeria showed that less than 40% of urban and 7% of rural households were willing to enrol both for themselves and other members of the households. However, in contrast, other reports such as that of Babatunde et al. (2012), Melaku et al. (2014), Bhawana et al. (2017) and Yusuf et al. (2019) showed a high level (87%, 77.8%, 71 and 65.8% respectively) of Willingness to enrol in a proposed health insurance scheme, while the study in Osun and the FCT by Bamidele et al. (2013) and Aderibigbe et al. (2017) respectively, showed that most of the respondents were willing to enrol, with most of them willing to make financial contributions in favour of themselves and family members. Also, the study of Bhawana et al. (2017) showed that the respondents had similar reasons for not enrolling for SHI (lack of finances and lack of believe in the authenticity of the scheme). Also related to this, trust was stressed in the studies of De Allegri (2006), Basaza et al. (2008) and Ozawa and Walker (2009), as very relevant to enrolment in any health insurance, while the main reasons for lack of willingness to enrol in the study of Tadele et al (2017) were identified as; limited health services given in SHI, perceiving poor quality services delivery and shortage of income

Also, among the few who expressed interest towards enrolment, majority stated that they would enrol in a private based scheme and the median range of contribution they are willing to make annually for their SHI was calculated as 70,000 Naira (5800 Naira monthly). This is higher than the amount reported in the study of Anderson & Adeniji (2019) where respondents were willing to pay N 676.96 ± 491.6 monthly. Other studies in Nigeria shows that household heads in Urban communities East and North are willing to pay ₦3,000 and ₦1,798.9 per person yearly while in the rural households in the North, it was found to be ₦721 (Usman & Bukola, 2013; Uzochukwu et al., 2015). The reason for the high amount is due to high earning power of the respondents in this region of the country.

### Factors associated with willingness to enrol in social health insurance

Analysis of the factors associated with the willingness of the respondents towards enrolling for SHI showed that age, educational level, employment status and monthly income are significantly associated (p<0.05) with willingness to enrol in SHI. Age in the study of Adebayo et al (2015) was also found to be significantly related to uptake of the scheme as younger individuals were more willing to pay compared to older individuals. In contrast, Shafie and Hassali (2013) showed that ethnicity and marital status are both associated with individuals’ willingness to pay, while Tadele et al. (2017) showed that age, sex, year of experience, educational status, and monthly salary did not show statistically significant association with willingness to pay for SHI.

Onwujekwe et al. (2010) reported that education and gender were the variables found to exert significant effects on willingness to enrol among respondents in their study in Southeastern region of Nigeria. This is also true of the studies conducted by Asenso-Okyere et al. (1997) and Dong et al. (2003) in Burkina Faso and Ghana respectively where sex is significantly associated to willingness to pay as male respondents were willing to pay a higher amount. Hence, Angel-Urdinola and Wodon (2010) concluded in their study that most household decisions in most African settings are usually affected by gender to the advantage of men and that men are usually the de facto head of household. Also, Adebayo et al. (2015) asserted that men are presumed to be responsible for financial decisions within the households in the African context.

The study by Anderson and Adeniji (2019) reported educational level, mode of payment, and marital status as statistically significant factors that influenced willingness to pay for SHI. Bamidele and Adebimpe (2012) also elicited that educational status of respondents was a key predictor of their practice towards health insurance in the Southwestern region of Nigeria. It was also reported that education plays a key role in uptake of health insurance (Adebayo et al., 2015), hence household heads with higher educational status in this study, were seen to be more willing to pay for SHI than those with lower educational status just as expressed in the studies of (Usman & Bukola, 2013; Uzochukwu et al., 2015). Furthermore, income or socioeconomic standing according to Wang et al. (2006) is very crucial in determining demand behaviour. Hence this study showed that employment status and monthly income are significantly associated with willingness to enrol in SHI. This is in line with other studies which showed that wealth or socioeconomic standing of households and individuals is associated with the willingness and ability to pay for health insurance (Kebede et al., 2014; Adebayo et al., 2015). As stated by Wang et al. (2006), the financial resource capacity of households and affordability of the premium are a first concern to enrol. Hence, those who are in good socio-economic level would be in a better position to enrol than their counter parts (Mirach et al., 2019).

## CONCLUSION

Data from this study has shown a huge number of the respondents are not enrolled in any healthcare insurance scheme, while over half of them indicated lack of interest in enrolling in SHI, with lack of trust in the management of the scheme, no believe in paying for sickness or have other means of meeting their healthcare needs identified as the major reasons behind this decision. Also, the main variables reiterated in the study as affecting the willingness of the respondents towards enrolling were identified as age, educational level, employment status and monthly. With respect to this finding, the study recommends health insurance companies should come out with clear policy details and carry out robust awareness campaigns to dispel false rumours about SHI and enlighten the public about its merits.

## Data Availability

All data produced in the present work are contained in the manuscript

